# Frequentmers - a novel way to look at metagenomic Next Generation Sequencing data and an application in detecting liver cirrhosis

**DOI:** 10.1101/2023.09.19.23295771

**Authors:** Ioannis Mouratidis, Nikol Chantzi, Umair Khan, Maxwell A. Konnaris, Candace S.Y Chan, Manvita Mareboina, Ilias Georgakopoulos-Soares

## Abstract

Early detection of human disease is associated with improved clinical outcomes. However, many diseases are often detected at an advanced, symptomatic stage where patients are past efficacious treatment periods and can result in less favorable outcomes. Therefore, methods that can accurately detect human disease at a presymptomatic stage are urgently needed. Here, we introduce “frequentmers”; short sequences that are specific and recurrently observed in either patient or healthy control samples, but not in both. We showcase the utility of frequentmers for the detection of liver cirrhosis using metagenomic Next Generation Sequencing data from stool samples of patients and controls. We develop classification models for the detection of liver cirrhosis and achieve an AUC score of 0.91 using ten-fold cross-validation. A small subset of 200 frequentmers can achieve comparable results in detecting liver cirrhosis. Finally, we identify the microbial organisms in liver cirrhosis samples, which are associated with the most predictive frequentmer biomarkers.

## Introduction

Early detection and diagnosis of human disease is essential for enhancing patient outcomes and reducing mortality rates (Lee, Huang, and Zelen 2004), as it enables timely efficacious intervention strategies. However, current modalities of detecting disease often lack the sensitivity and specificity required to capture presymptomatic stages of disease progression accurately. Therefore, there is an urgent need to establish novel methods that identify unique biological markers of disease which precede the manifestation of symptoms.

Sequencing has provided the opportunity to investigate the molecular insights of disease mechanisms and potentially identify unique changes between healthy and affected individuals. Kmers, contiguous DNA subsequences of length *k*, have been successfully implemented across multiple research problems including in the construction of genome alignments (Rahman et al. 2018), for the generation of genome assemblies (Rhie et al. 2020), in understanding evolutionary relationships between species (Yang et al. 2020) and the construction of phylogenies (Bussi, Kapon, and Reich 2021) among other applications. Additionally, a number of algorithms have been developed for the faster and more efficient derivation of kmers and their occurrences, such as Jellyfish (Marçais and Kingsford 2011) and BBDuk (Bushnell, Rood, and Singer 2017). Kmers have been previously used to describe new features or characteristics of an organism related to the presence or absence of a specific contiguous subsequence. For example, the subset of kmers that do not appear in a genome are referred to as nullomers (Acquisti et al. 2007; Georgakopoulos-Soares, Yizhar-Barnea, et al. 2021; Koulouras and Frith 2021) and the subset of kmers that are found in a single species are referred to as quasi-primes (Mouratidis et al. 2023). Using kmer strategies, we may efficiently mine the human genome for differences that distinguish patients with disease from healthy individuals in effort to establish unique biological signatures.

Liver cirrhosis is a major health burden across countries, affecting 5.2 million people globally and causing 1.48 million deaths in 2019 alone (Y.-B. Liu and Chen 2022). The proportion of liver cirrhosis deaths as a fraction of total deaths in the population has increased in the last decade, indicating the need for early detection and intervention, including lifestyle changes and treatments (GBD 2017 Cirrhosis Collaborators 2020). Metagenomic Next Generation Sequencing (mNGS) is a powerful tool that enables researchers and clinicians to identify and characterize microbial pathogens, antimicrobial resistance, and virulence markers from various samples, which can facilitate early disease detection and diagnosis. In a study conducted by Qin et al. (Qin et al. 2014), a cohort of 123 patients with liver cirrhosis and 114 healthy individuals was studied using mNGS data from stool samples (Qin et al. 2014). Qin et al. trained a disease classifier using leave-one-out cross-validation on 98 patient and 83 healthy control samples to identify gene markers enriched either in patients or controls. The computational complexity of this cross-validation approach informed the decision of the authors to only use fifteen gene markers as features for the Support Vector Machine (SVM) model, achieving an AUC value of 0.836. Improvements in the performance of such a model would be required for the clinical implementation of mNGS for liver cirrhosis detection.

Here we describe a feature selection approach for machine learning models based on a novel feature termed frequentmers. We define frequentmers as kmers that are present in multiple samples from one group but completely absent from the other group. We extracted frequentmers across the mNGS dataset from the liver cirrhosis study, utilizing them to train a machine learning model that achieves an AUC of 0.91 with 10-fold cross-validation. We demonstrated that a small number of 200 frequentmers can result in comparable classification accuracy. We also identified specific microbial species that are informative for the detection of liver cirrhosis. Frequentmers are transferable to other diseases and sequencing assays, representing a novel method for biomarker development and for the detection of human diseases.

## Results

### Derivation of frequentmers

We have derived a new type of algorithm that identifies highly informative and specific sequences to enable the early detection of human diseases (**Figure 1**). First, we identified the set of kmer sequences that are observed in each patient and each control sample. Next, we calculated the number of samples in which each kmer sequence is present. We removed sequences that are present in both patient and control samples, hypothesizing that these sequences are less likely to reflect differences between the two groups, and to serve as biomarkers. The subset of sequences that are found in multiple healthy control samples and not in patient samples is termed “control frequentmers” and similarly the subset present in multiple patient samples and absent from healthy control samples is termed “patient frequentmers”. The aforementioned process is performed using ten-fold cross-validation. For each fold, 90% of the samples are used as a training set and the remaining 10% is used as a test set. Frequentmers are derived independently from the training set of each fold. The mathematical formulation is provided below.

**Figure 1:**
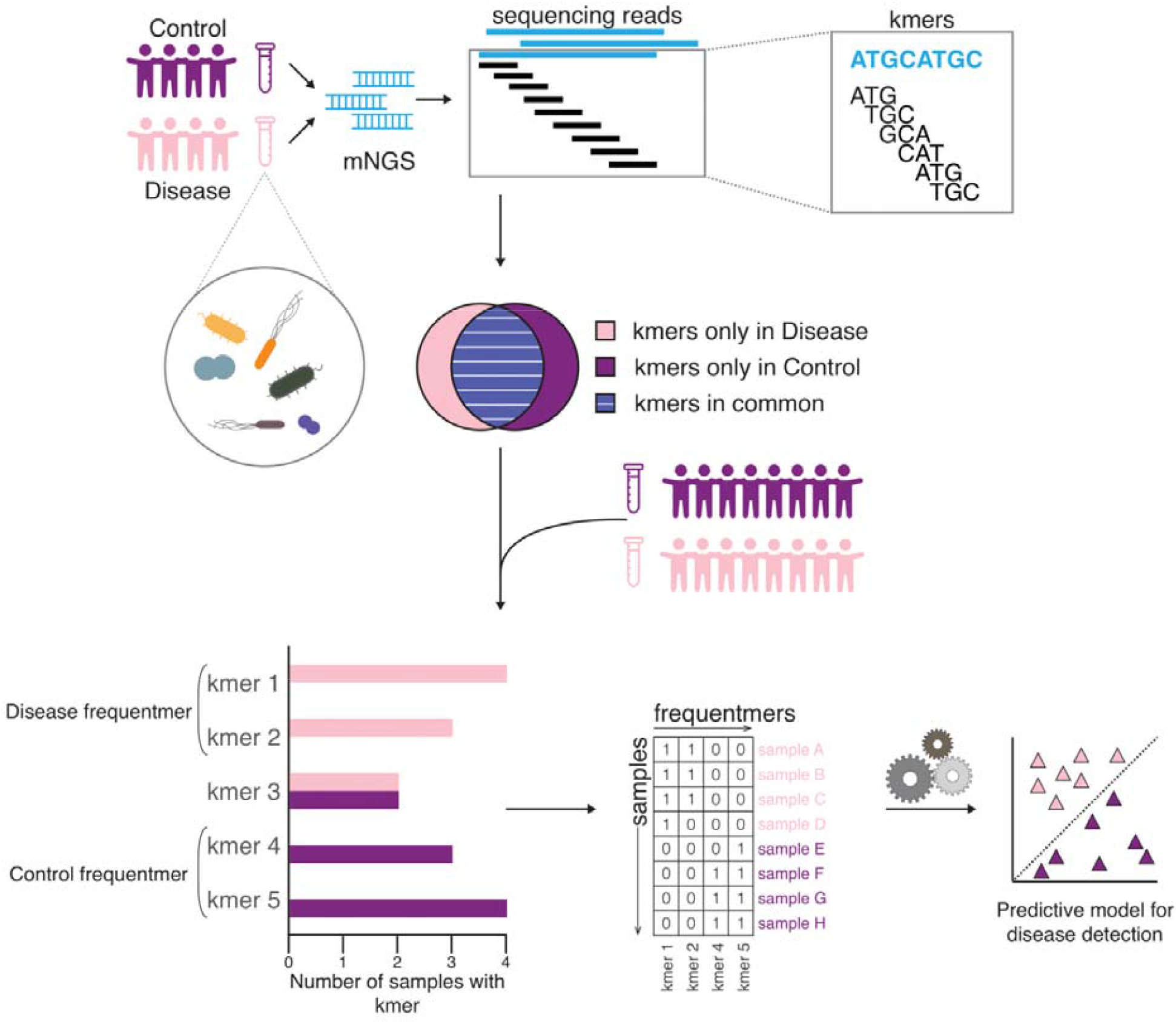
Visualization of frequenter extraction pipeline and inference. Two groups of samples are examined, the first group is composed of healthy control samples and serves as the control and the second group contains patient samples, for the disease that is investigated. mNGS data are analyzed to determine the number of kmers found in each sample and subsequently the kmers unique to only one group (healthy controls or patients) are identified. Frequentmers represent the recurrent kmers found only in patient samples or only in healthy control samples, but never in both. Frequentmers that are found in multiple samples of only one group are used as features to train a machine learning algorithm to perform binary classification on unseen data.

### Definitions

Let us define alphabet *L = {A, T, C, G}* representing adenine, thymine, cytosine, and guanine respectively. Metagenomic Next Generation Sequencing reads can be represented as a nucleotide string *R = t*_*1*_*t*_*2*_*t*_*3*_… *t*_*z*_ over this alphabet. We can then represent the entirety of an individual’s sequenced metagenome as a collection of strings *I = {R*_*1*_, *R*_*2*_, …, *R*_*n*_*}*.

A nucleotide kmer *K* is defined as a short nucleotide sequence of length k over alphabet L and can be represented as *K = s*_*1*_*s*_*2*_ *s*_*3*_ … *s*_*k*_. A kmer *K* is said to belong to a read *R, k* ∈ *R*_*i*_, if and only if ∃*i, j* ∈ {1, …, *z*}: *j* – *i* = *k* - 1 ∧ *s*_*1*_*s*_*2*_ *s*_*3*_ … *s*_*k*_= *t*_*i*_*t*_*2*_*t*_*3*_ … *t*_*j*_. Note that *j - i= k - 1* implies the kmer is comprised of exactly *k* nucleotides.

A kmer *K* is said to belong to an individuals metagenome *I* if and only if ∃i: *k* ∈ *Ri* ∧ *Ri* ∈ *I*.

The samples used for training our algorithm can further be subdivided into two distinct groups, mNGS sequencing of samples taken from healthy control samples and mNGS sequencing taken from individuals with liver cirrhosis. Let us name the two groups *H* = *{H*_*1*_, *H*_*2*_, *H*_*3*_,…, *H*_*m*_ *}* and *P = {P*_*1*_, *P*_*2*_, *P*_*3*_, …, *P*_*i*_ *}* of controls and patients respectively.

A kmer *K* is said to be a healthy control frequentmer of recurrency *r* if and only if:

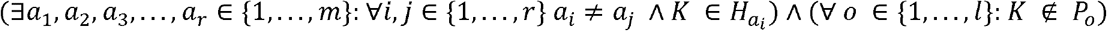

In other words, a kmer *K* is said to be a healthy control frequentmer of recurrency *r* if and only if this k-mer appears in at least r control samples and does not appear in any patient samples.

Similarly, a kmer *K* is said to be a patient frequentmer of recurrency *r* if and only if:

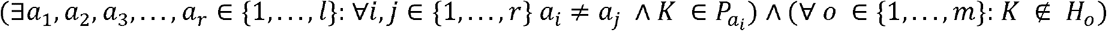

In other words, a kmer *K* is said to be a patient frequentmer of recurrency *r* if and only if this kmer appears in at least *r* patient samples and does not appear in any control samples.

### A disproportionate number of liver cirrhosis-specific kmers is detected

We implemented our algorithm in metagenomic Next Generation Sequencing (mNGS) data derived from fecal samples of liver cirrhosis patients and healthy controls (Qin et al. 2014). In total, we examined 123 patients with liver cirrhosis and 114 matched healthy controls. We extracted every sixteen base-pair (bp) kmer found in each sample and split samples in ten groups or folds, with the proportion of cases over the total samples in each fold being maintained. The choice of kmer length was informed from previous studies in which we found that the performance of the kmer-based models increased as a function of kmer length up to sixteen bp length (Georgakopoulos-Soares, Barnea, et al. 2021). For each fold, we examined which subset of the total kmers detected constituted frequentmers, using the number of samples in which each kmer was found as the recurrency threshold (see Methods). Thus, we estimated the number of healthy control and patient frequentmers as a function of the recurrence among liver cirrhosis patients and healthy controls, respectively.

First, we find that the number of frequentmers recovered decreases as a function of the recurrency threshold used (**Supplementary Figure 1a-c)**. We also find that as the recurrency threshold for the number of samples a frequentmer is present in increases, there is a larger proportion of the total frequentmers being patient frequentmers relative to control frequentmers (**Figure 2a-b; Supplementary Figure 2;** Pearson correlation: r = 0.975, p-value<e-9). Specifically, we observe that for the recurrency threshold of five samples, there is 8.92-fold more patient than healthy control frequentmers, whereas at recurrence of twenty samples there is 306.5-fold more patient than healthy control frequentmers (**Figure 2b;** binomial test, p-value = 0), indicating an imbalance between the number of healthy control and patient frequentmers identified. These differences likely stem from changes in the microbiome of liver cirrhosis patients, which are observed recurrently across multiple liver cirrhosis patients and which are not normally observed in healthy microbiomes.

**Figure 2:**
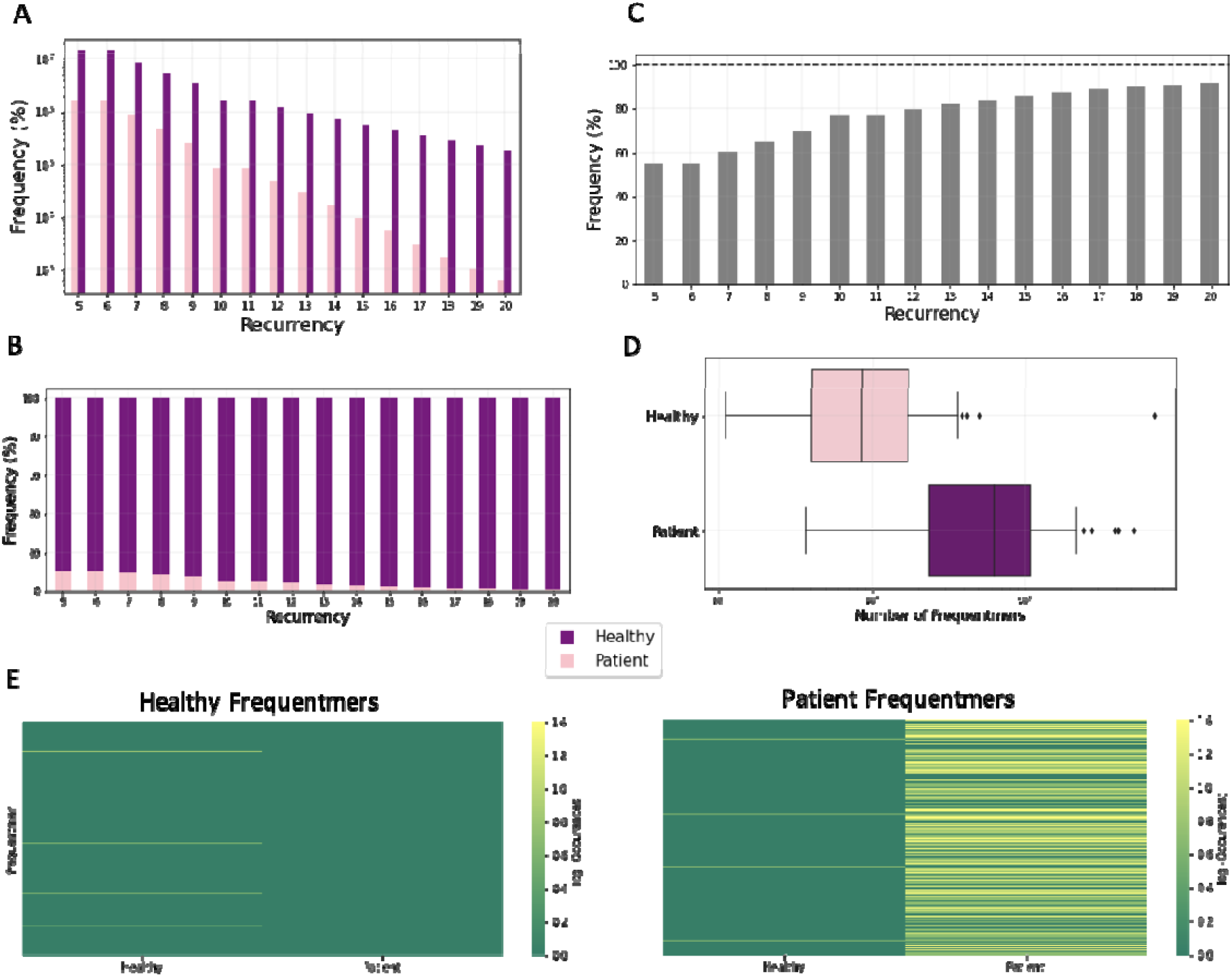
Characterization of frequentmers associated with liver cirrhosis. **A**. The number of liver cirrhosis frequentmers and healthy control frequentmers identified as a function of the number of samples in which they were detected (recurrency). **B**. Stacked barplot showing the proportion of the total frequentmers being patient and healthy control frequentmers. **C**. Bar plot displaying the frequency with which frequentmers identified in the training set are observed in the test set, for recurrency thresholds 5-20. Results shown represent the mean across the ten folds. **D**. Number of frequentmers detected in the test set for healthy control and liver cirrhosis frequentmers for recurrency threshold of fifteen across folds. Results shown represent the mean across the ten folds. **E**. Number of healthy frequentmers in the training set also detected in the test set of healthy control and patient samples (left). Number of liver cirrhosis frequentmers in the training set also detected in the test set of healthy control and patient samples (right). Frequentmers of the recurrency threshold of fifteen samples were used.

Next, we examined what proportion of frequentmers observed in the training cohort was also identifiable in the test cohort. We find that the proportion of frequentmers observed in the test cohort is correlated with the recurrence threshold (**Figure 2c; Supplementary Figure 3;** Pearson correlation: r = 0.964, p-value<2.03e-9). We also observe that for recurrence threshold of fifteen samples, 86% of frequentmers are recovered, with the proportion of frequentmers that is recovered leveling off around this recurrence threshold (**Figure 2c)**. Importantly, the number of patient frequentmers detected in the test set is significantly larger in samples from liver cirrhosis patients relative to healthy controls across the recurrency thresholds examined (**Figure 2d;** Mann-Whitney U, p-value=0). Additionally, we find that healthy control frequentmers from the training set are 6.49-fold more likely to be found in healthy control samples in the test set (**Figure 2e;** Mann-Whitney U, p-value<0.00016). Similarly, liver cirrhosis frequentmers derived in the training set are 9.04-fold more likely to be found in liver cirrhosis samples in the test set (**Figure 2e;** Mann-Whitney U, p-value<9.1e-5), providing further support for efficacy of our methodology. Therefore, we find that across the ten folds that were independently evaluated, there are frequentmers that are consistently detected and that are either liver-cirrhosis specific or only derived from healthy control samples.

**Figure 3:**
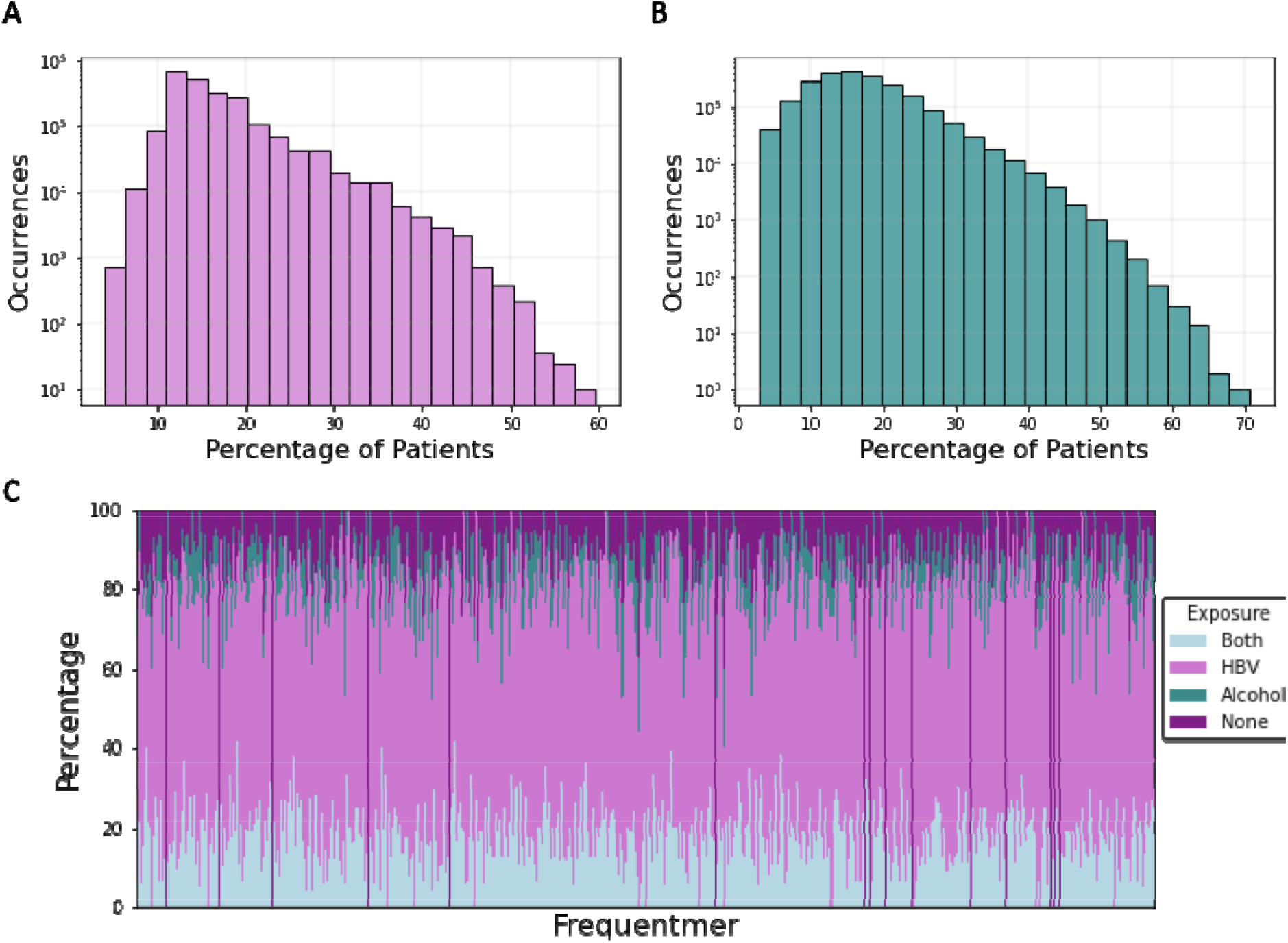
Characterization of liver cirrhosis frequentmers in relationship to HBV infection and alcohol consumption. **A**. Histogram displaying the number of frequentmers and the corresponding proportion of HBV-positive patient samples they were detected. **B**. Histogram displaying the number of frequentmers and the corresponding proportion of alcohol samples in which they were detected. **C**. Frequentmer distribution in samples that are HBV-positive (n=99), have high alcohol intake (n=34), are both HBV-positive and have high alcohol intake (n=23) or are not associated with either (n=13).

### Identification of kmers associated with HBV infection and high alcoholic consumption

Liver cirrhosis is linked to both high alcohol intake and HBV infection (Scaglione et al. 2015). For liver cirrhosis, we investigated if there were differences in the samples that are Hepatitis B virus (HBV) positive, regarding the frequentmers detected.

Among the liver cirrhosis frequentmers of recurrency fifteen, we find that 55,789 are specific to liver cirrhosis patients that are HBV positive and are completely absent from HBV negative patients, for recurrency (**Figure 3a; Supplementary Figure 3**). Similarly, we examined samples from alcohol-related liver cirrhosis patients and observed that 5,004 are specific to them for recurrency threshold of five (**Figure 3b; Supplementary Figure 4**). The recurrency threshold of five and fifteen were selected as they represent an approximately equal proportion of the total number of samples that are alcohol-related and HBV positive, which are 34 and 99 respectively. Therefore, we conclude that we can detect healthy control frequentmers, general liver cirrhosis frequentmers and frequentmers that reflect viral exposure (HBV) and lifestyle (alcohol consumption) differences. We find that the majority of frequentmers originate primarily from HBV positive samples, which is to be expected as they constitute the majority of our patient samples. However, there are differences in the distribution of frequentmers, with some tending to be found more in either HBV positive or alcohol-related samples (**Figure 3c**).

**Figure 4:**
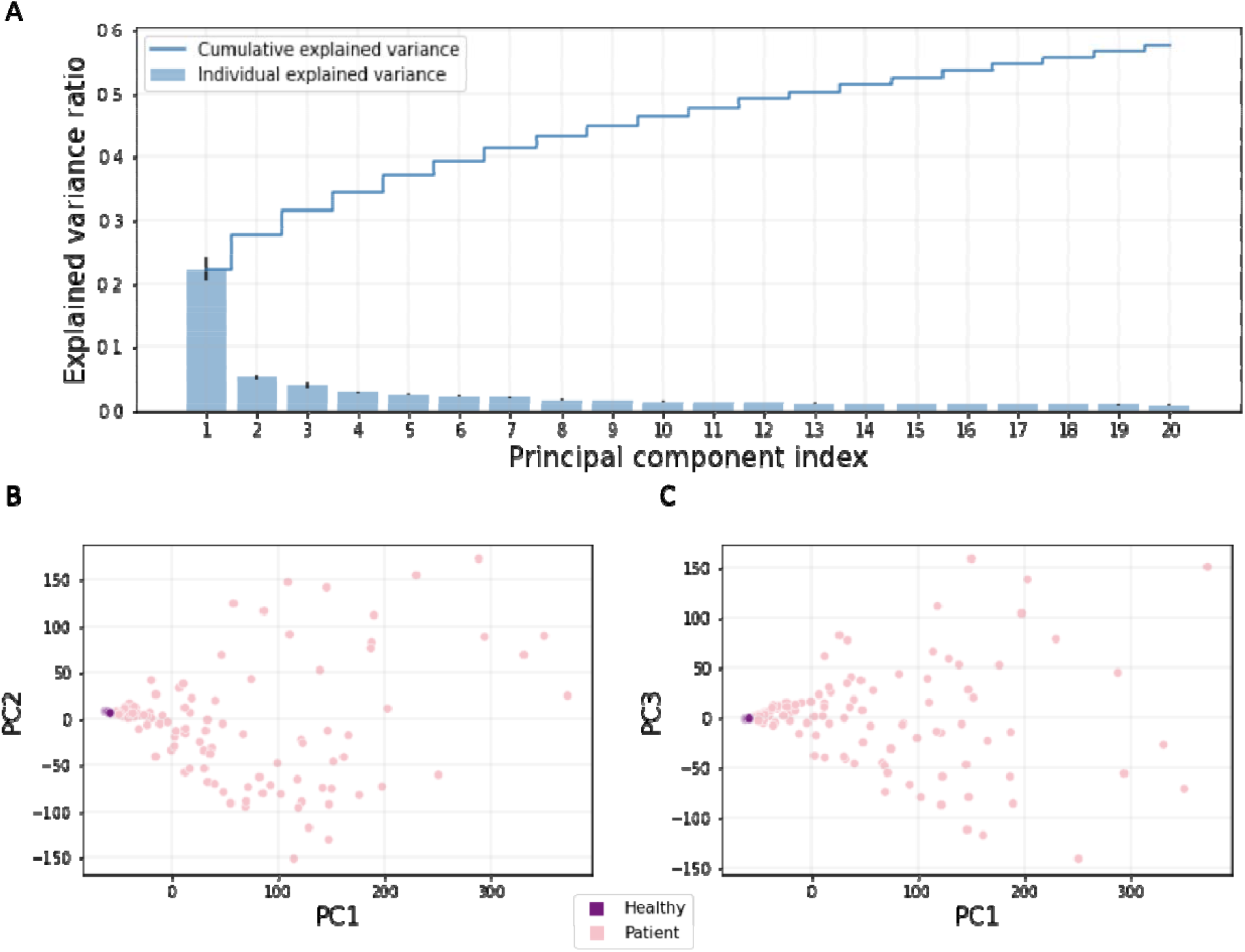
**A.** Proportion of variance explained by twenty principal components. Mean score of the explained variance ratio across the ten folds is shown. Error bars show standard deviation. Line plot indicates the cumulative explained variance across the twenty first principal components. **B-C**. Scatter plot displaying the separation of patient and control samples by the first three principal components. Results shown for **B**. PC1 versus PC2 and **C**. PC1 versus PC3.

We examined the number of frequentmers of recurrency fifteen shared in HBV positive patients relative to patients that were HBV negative and observed that the two groups were dissimilar in their frequentmer profile (Mann-Whitney U, p-value<0.0143). Similar results were observed for samples that were derived from high alcohol intake patients relative to other patients (Mann-Whitney U, p-value<3.49e-6). We conclude that differences in the exposures of samples are reflected in their frequentmer profile.

### Principal Component Analysis reflects differences in frequenter profiles

Next, we examined if liver cirrhosis and healthy control samples are linearly separable. A principal component analysis (PCA) was used to examine the information that healthy control and liver cirrhosis frequentmers can capture to separate samples from the two groups. We observe that a large fraction of the variance can be explained by the first twenty principal components (PCs), with the first PC alone capturing 22.46% of the variance (**Figure 4a**). Additionally, we observe that the first three PCs can separate the liver cirrhosis and control samples (**Figure 4b-c**). These findings provide evidence that frequentmers can capture differences in the mNGS profile of liver cirrhosis patients and healthy controls.

### A predictive model based on frequentmers can accurately detect liver cirrhosis

The early detection of liver cirrhosis is critical for intervention and improved clinical outcomes of patients (Trivedi and Tapper 2018). We therefore developed machine learning classification models to examine if frequentments can accurately predict liver cirrhosis patients from healthy controls. The first model we examined was a logistic regression model, which has inherent advantages such as interpretability and determinism. We examined the performance of the model using multiple recurrency thresholds for the number of samples in which each frequentmer was found in the training set. We observed that when increasing the sample recurrency threshold, the performance of the model increased (**Supplementary Figure 6**), which is likely due to removing features that were less informative. We also report that the logistic model has an AUC of 0.91 for recurrency threshold of fifteen samples (**Figure 5a-b**), indicating that it can accurately detect liver cirrhosis. The performance of our model was superior to that obtained from the original article (Qin et al. 2014). We also find that the top features are liver cirrhosis frequentmers (**Supplementary Figure 7-8**). From the 1,000 most informative coefficients (as measured by absolute coefficient score) of the logistic regression model 993 were liver cirrhosis frequentmers, which was significantly more than expected by chance (Binomial test, p-value<1.4e-07; **Figure 5c, Supplementary Figure 7)**. As a result, we conclude that our feature selection is largely reflected in what the logistic regression model learns and is primarily based on liver cirrhosis frequentmers.

**Figure 5:**
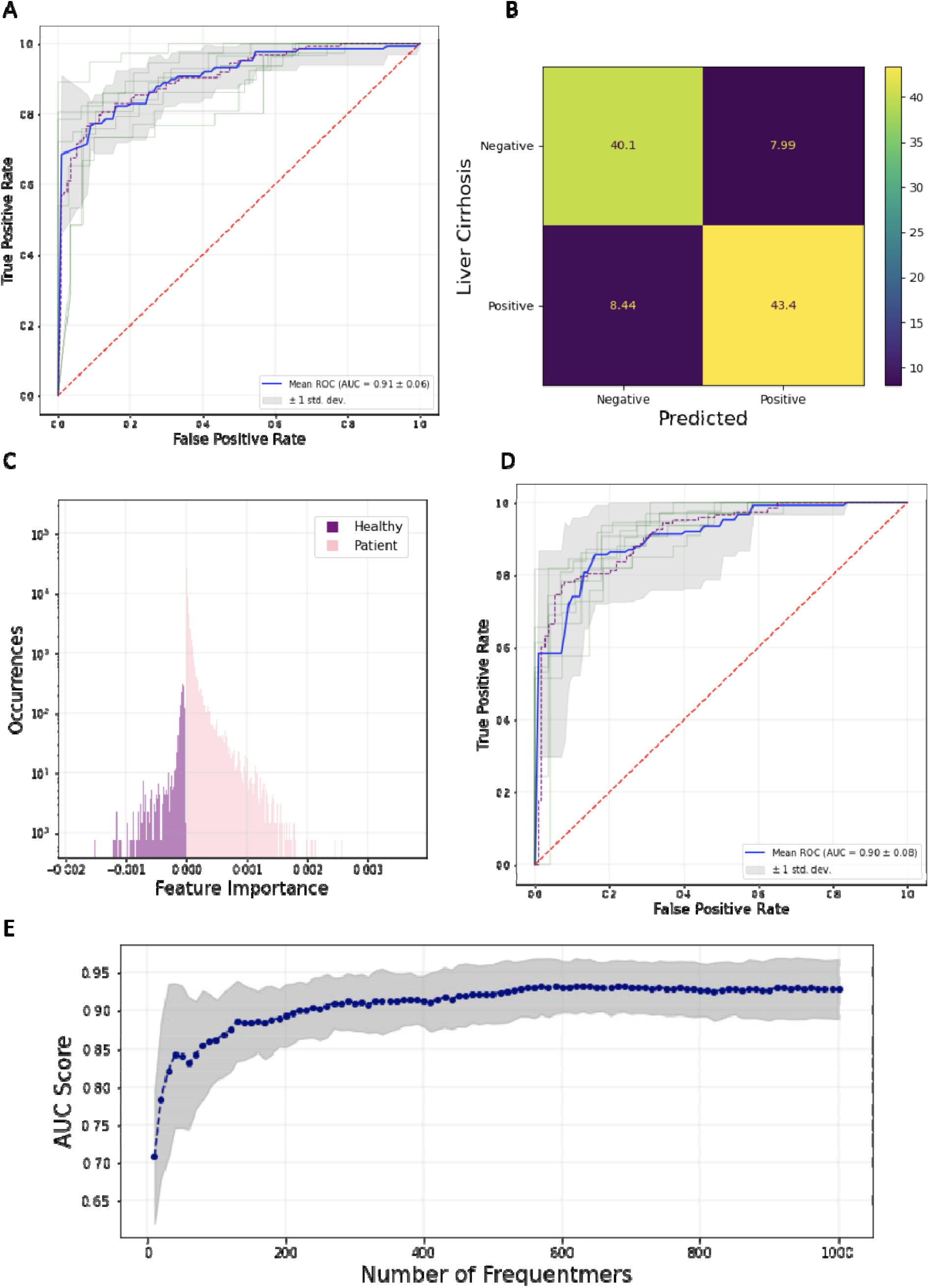
Machine learning based liver cirrhosis detection. **A**. ROC curve displaying the AUC for the logistic regression model for recurrency threshold of fifteen. **B**. Confusion matrix showing the percentage of samples that were correctly and incorrectly classified as liver cirrhosis patients or healthy controls, for recurrency threshold of fifteen. **C**. Logistic regression classification coefficients. **D**. ROC curve displaying the AUC for the XGBoost classification model, for recurrency threshold of fifteen. **E**. AUC score relative to number of top frequentmers used for logistic regression. Gray lines display the confidence intervals from the ten folds. The blue line shows the mean AUC score across the ten folds.

**Figure 6:**
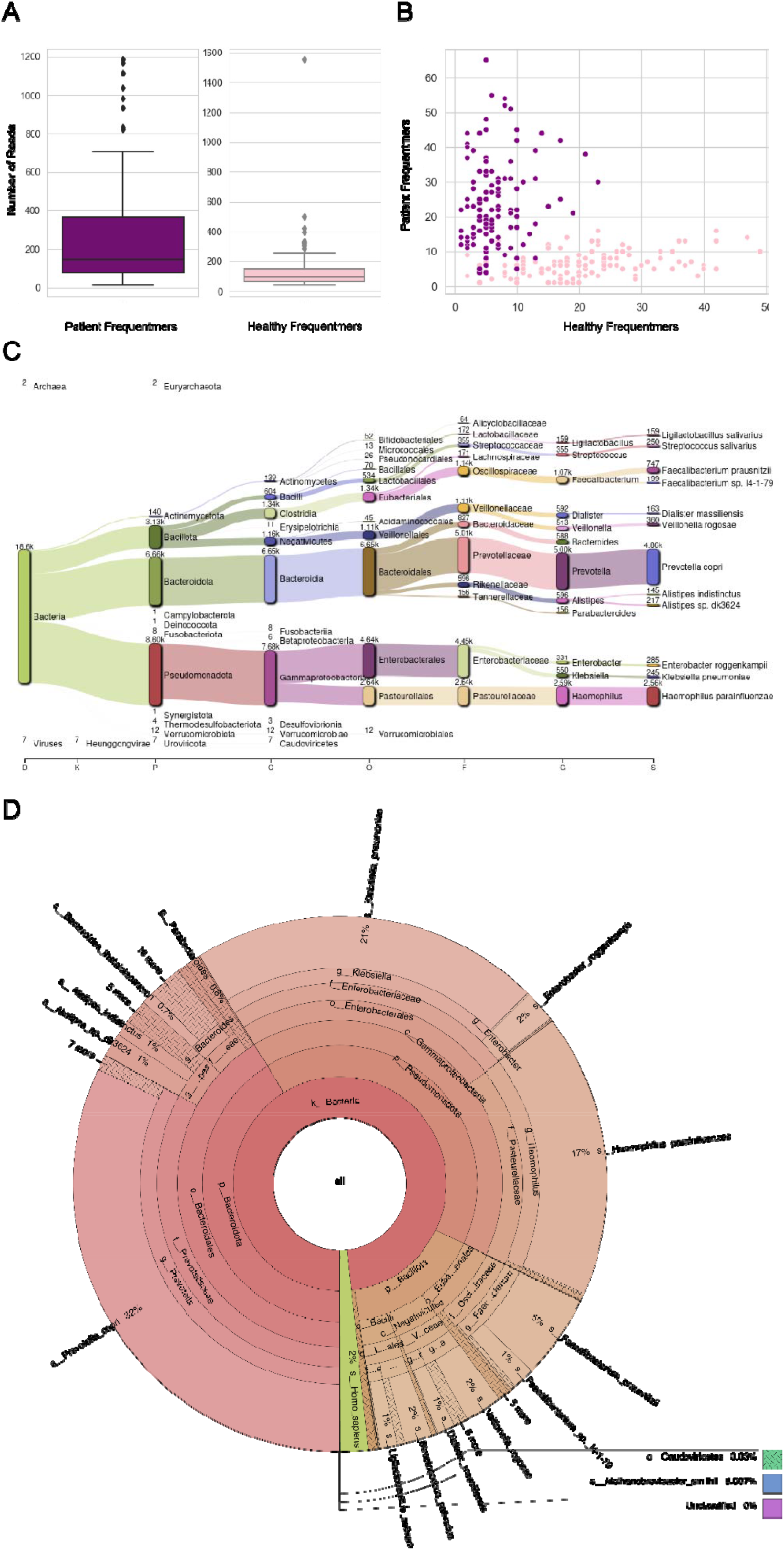
Frequentmer containing reads. Top and bottom 100 most informative frequentmers based on logistic regression with recurrency threshold fifteen. **A**. The corresponding number of reads originated from. **B**. Their distribution across patient and healthy control samples. **C**. Taxonomic assignment for the top frequentmer reads across microbial organisms. **D**. Krona Radial space-filling plot showing the identified microbial species abundance.

To examine non-linear patterns between frequentmers we also implemented an XGBoost classification model. Extreme Gradient Boosting is a classification framework based on training a sequence of decision trees and utilizing their combined predictions to make the final classification (T. Chen and Guestrin 2016). We observe that across the different sample recurrency thresholds the model performs comparably to the logistic regression model (**Supplementary Figure 9, Figure 5d**). At low recurrency thresholds, for which the number of features is extremely large and the noise in the system increases, XGBoost outperforms logistic regression (**Supplementary Figure 6, Supplementary Figure 9**). However, for a frequentmer recurrency threshold of fifteen samples we obtained an AUC score of 0.90, suggesting that the ensemble model did not perform better than the logistic regression model.

Additionally, we were interested to find if we can perform similarly in detecting liver cirrhosis using only a small fraction of the frequentmers. We therefore investigated how the number of frequentments used in the classification model influenced the performance. To achieve this we identified the most informative features from the training set of each fold in the logistic regression using the absolute value of each logistic regression coefficient for each frequentmer (**Figure 5c**) and re-trained the logistic regression model for the same samples in the training set. We then tested the performance of our model. This process was repeated, examining between 25 and 1,000 frequentmers. We observe that even with roughly 200 frequentmers we achieve comparable performance to the original model that used all frequentmers (**Figure 5a, e**). These results indicate that with a small number of frequentmers we can be used to generate a classification model that can accurately detect liver cirrhosis.

### Identification of microbial species driving the classification models

Utilizing the coefficients of the logistic regression performed at recurrency threshold fifteen, which corresponded to the best performing model, we identified the 100 frequentmers with the highest positive regression coefficient and the 100 frequentmers with the lowest negative regression coefficient averaged over all ten folds. Within this group, the 100 frequentmers with a positive coefficient were patient frequentmers and the 100 frequentmers with a negative coefficient were healthy control frequentmers.

We then extracted the sequencing reads from which those frequentmers originated, identifying a total of 41,944 reads. On average, these frequentmers were present in 210 reads (mean: 209.72, standard deviation: 256.50), with some significant outliers skewing the variance (**Figure 6a**). Patient frequentmers were found in significantly more reads than healthy frequentmers (t-test, p-value X). The distribution of samples and the number of patient and healthy frequentmers supports a clear separation between patient and control samples (**Figure 6b**).

We further identified the microbial species from which the frequentmers were derived from. Interestingly, we find a set of bacterial species that are highly enriched for frequentmers (**Figure 6c-d**), including *Prevotella copri, Haemophilus parainfluenzae, Faecalibacterium prausnitzii* and *Klebsiella pneumoniae*, multiple of which have been previously associated with liver cirrhosis (Dong et al. 2020) or other liver-associated diseases such as liver abscess (Y. Liu, Wang, and Jiang 2013), and Nonalcoholic Fatty Liver Disease (Hu et al. 2022). Therefore, we conclude that we can identify the microbial species from which the frequentmers were derived, showcasing the interpretability of our approach.

## Discussion

In this work, we describe the development of a method that enables the identification of kmer sequences that are specific to patient and healthy control samples, which we term patient and healthy control frequentmers, respectively. We show that frequentmers can be used as disease detection biomarkers, by demonstrating their utility for the detection of liver cirrhosis using mNGS data, in which we outperform previously published models and achieve an AUC score of 0.91 (Qin et al. 2014).

The integration of other biomarkers, clinical information and risk factors can result in further improvements of our models for the early detection of human disease. Furthermore, a major strength of our method is the interpretability of our logistic regression model and we provide evidence that we can directly infer the microbial species from which the frequentmers are derived from. We show that the majority of high importance frequentmers originate from microorganisms known to be associated with liver cirrhosis, such as *Prevotella copri, Faecalibacterium prausnitzii* and *Klebsiella pneumoniae*, multiple of which have been previously associated with liver cirrhosis (Dong et al. 2020; Y. Chen et al. 2021; Yuan et al. 2019). These results demonstrate the ability of the method to discover new associations between microbial species in the gut microbiome and disease. Investigation of the biological function of these microbial species and their roles in liver damage and cirrhosis are of particular interest for future work. Therefore, frequentmers can provide insights into microbial changes specific to the development of liver cirrhosis which could result in mechanistic insights for the role of the microbiome in this disease.

Examination of fecal samples using mNGS data is a non-invasive procedure that can be used for the early detection of liver fibrosis, before the manifestation of symptoms associated with liver cirrhosis. A small set of frequentmers suffices to achieve high predictive power in detecting liver cirrhosis (**Figure 5e**), which is another important advantage of our method. As a result, the detection of a small set of frequentmers could enable novel diagnostics based on short DNA sequences from mNGS data. For instance, CRISPR-based detection tools could be used to target frequentmers in detection assays (Kellner et al. 2019) or sequencing-based approaches such as adaptive sampling (also known as selective sequencing), which can enrich for specific sequences (Loose, Malla, and Stout 2016), can be applied to reduce detection costs.

A number of human diseases are associated with changes in the human microbiome, including cancer (Helmink et al. 2019) neurodegenerative diseases (Romano et al. 2021), metabolic diseases (Fan and Pedersen 2021), autoimmune disorders (De Luca and Shoenfeld 2019; Franzosa et al. 2019) and various infections (Whiteside et al. 2015; Natalini, Singh, and Segal 2023; Honda and Littman 2012). Therefore, machine learning models based on frequentmers could be applicable towards the detection of multiple other diseases as well as for pathogen detection. Furthermore, our methodology can be transferred across different experimental assays beyond mNGS, and it will be of interest to investigate its utility for cfDNA and cfRNA based diagnostics. The origin of the biological material can also vary and in future work it will be of interest to develop frequentmer based classification models for urine, saliva and blood samples.

In summary, we provide a novel methodology for the derivation of disease detection biomarkers and showcase their utility in the detection of liver cirrhosis from mNGS data obtained from fecal samples. Future work is required to analyze additional datasets and expand our findings in a multi-disease detection assay that is based on disease-specific frequentmers.

## Methods

### Retrieval and preprocessing of mNGS data

mNGS data from fecal samples of 123 liver cirrhosis patient samples and 114 healthy control samples were derived from (Qin et al. 2014). Across all samples, sequencing reads were examined as single-end. For samples with multiple sequencing runs, the sequencing reads across the runs were merged.

### Train-test split

In order to properly validate our results given our limited number of samples we performed ten-fold cross-validation. To that effect, we created ten different folds assigning 90% of the samples to the training and 10% in the test sets. Each fold consisted of liver cirrhosis patients and healthy control samples.

### Identification of kmers in each sample

For each sample, kmers of sixteen bp length were extracted using the Jellyfish package (Marçais and Kingsford 2011). If a kmer appeared only once in a sample it was discarded from downstream sequencing analysis as a potential sequencing error.

### Derivation of frequentmers

We defined two groups, the first consisted of only healthy control samples and the second consisted only of liver cirrhosis patient samples. Frequentmers of recurrency r were defined as kmers that appeared in a minimum of r samples of the same group and were absent from every sample of the other group. To avoid over-fitting, the extraction of frequentmers was performed for each fold separately.

Identification of HBV or high alcohol consumption associated frequentmers was performed by analyzing the frequentmers present in HBV-positive samples or samples of individuals with high alcohol consumption.

To measure if the frequentmer profile between patient samples that were HBV positive or had high alcoholic intake differed from other patient samples we estimated the jaccard index based on the number of shared frequentmers between patient samples and examined the jaccard index distribution across all pairs in the two groups using paired t-tests.

### Frequentmer analysis

Recurrency thresholds of zero, three, five, ten and fifteen samples were examined. Analyses were performed independently for each recurrency threshold. For each recurrency threshold, results were averaged across the ten folds.

Majority voting, if it was found in more healthy control samples or more liver cirrhosis samples in the test set, was used to classify frequentmers found in the test set and calculate Mann-Whitney U statistic.

### Principal Component Analysis

To examine if frequentmers can linearly separate the liver cirrhosis patient samples from the healthy control samples we implemented principal Component Analysis with 90 components. The first three principal components were used to visually inspect the separation of the patient and the control samples. Principal Component Analysis was performed with the scikit-learn package (Pedregosa et al., n.d.).

### Classification models

Logistic regression was performed using healthy control and liver cirrhosis frequentmers as features with the scikit-learn package, using the parameters: penalty: Ridge (L2), max_iter: 2000 and C (the inverse regularization strength): 0.01. For each frequentmer, the coefficient score was derived and distribution histograms were generated for healthy control and liver cirrhosis frequentmers separately. The XGB-boost classification model was generated using the package from https://github.com/dmlc/xgboost with the parameters: max_depth=11, gamma=0.3, eta=0.2, alpha=6 (T. Chen and Guestrin 2016).

To examine how the number of frequentmers used to train the logistic regression model affected the performance of the model, we used the absolute value of the logistic regression coefficients in the training set to re-train a model with the same sample split into training and test sets. The number of features examined ranged between 25 and 1,000 and performance was measured with the AUC score of each model. This process was repeated separately for each fold from which we derived the mean AUC score and confidence intervals across the ten folds.

### Frequentmer identification in microorganisms

Kraken2 taxonomic classification (Wood, Lu, and Langmead 2019) using the standard reference database was performed for the reads containing the frequentmers with the highest and lowest coefficients from the logistic regression model. The standard reference database was built using adjusted parameters --kmer-len 16 --minimizer-len 15 --minimizer-spaces 3. An alluvial plot/sankey diagram was generated using Pavian (Breitwieser and Salzberg 2020). Using the taxonomy labels generated from Kraken2, Bracken was performed to produce estimates of species- and genus-level abundance of each species (Lu et al. 2022). The KrakenTools suite was used to calculate statistics and format the output from Bracken for visualization with Krona (Ondov, Bergman, and Phillippy 2011). Krona was used to generate an RSF display that visualizes the Bracken output of the species- and genus-level relative abundance.

## Data Availability

The analyzed during the current study are available in the European Nucleotide Archive repository, PRJEB6337.

## Declarations

### Ethics approval and consent to participate

Not applicable

### Consent for publication

Not applicable

### Availability of data and materials

All the associated code used for the generation of figures and presentation of data throughout the manuscript is deposited on GitHub at the following link: https://github.com/Georgakopoulos-Soares-lab/frequentmer_analysis

### Author contributions

I.M. and I.G.S., conceived the study. I.M., N.C., M.A.K., M.M., and I.G.S., wrote the code, I.M., N.C., M.A.K., C.C., and I.G.S., performed the analyses and generated the visualizations. I.G.S. supervised the research. I.M., N.C., M.A.K., and I.G.S. wrote the manuscript with input from all authors.

## Acknowledgements

I.M., N.C., M.K., and I.G.S. were funded by the startup funds from the Penn State College of Medicine. We would like to thank Martin Hemberg for offering useful comments.

