## Supplemental Figures for "Frequentmers - a novel way to look at metagenomic Next Generation Sequencing data and an application in detecting liver cirrhosis"

**Supplementary Figures**

A

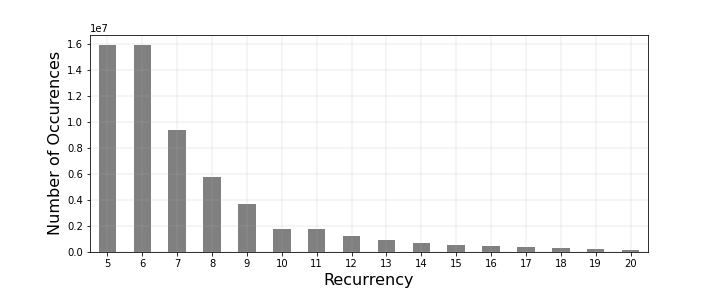

B

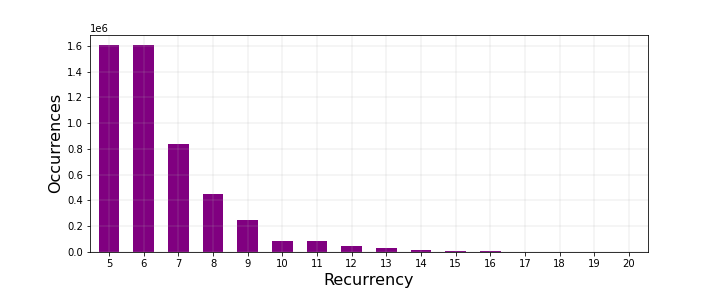

C

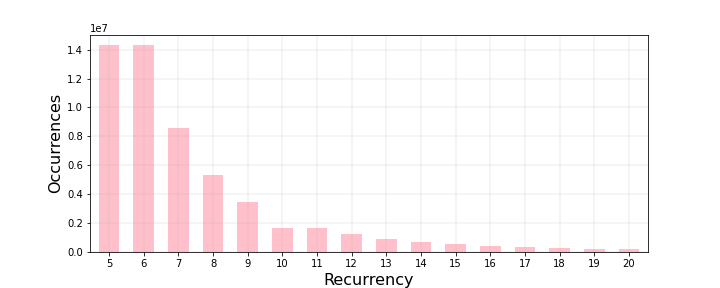

**Supplementary Figure 1: Number of frequentmers detected as a function of the recurrency threshold.** Recurrency thresholds of five to twenty samples were examined. Results shown for: **A.** healthy control and liver cirrhosis frequentmers, **B.** healthy control frequentmers, **C.** liver cirrhosis frequentmers.

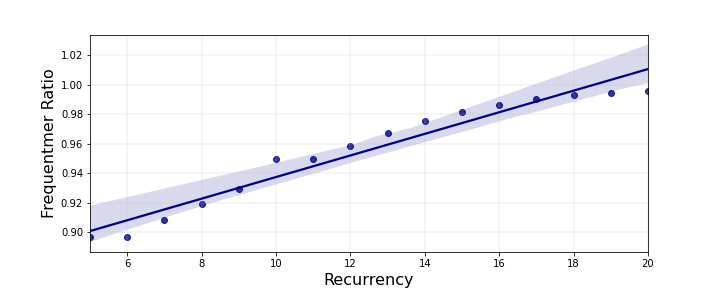

**Supplementary Figure 2: As the recurrency threshold increases a larger proportion of frequentmers are patient frequentmers.** Frequentmer ratio was defined as the ratio of patient frequentmers over healthy control and patient frequentmers. Values are averaged over ten folds. 99th percentile confidence intervals are shown.

A	B

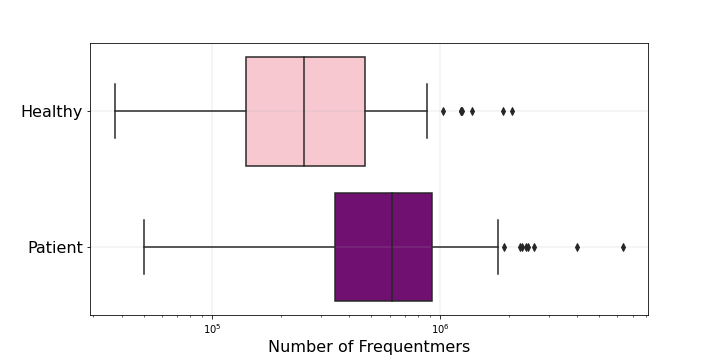

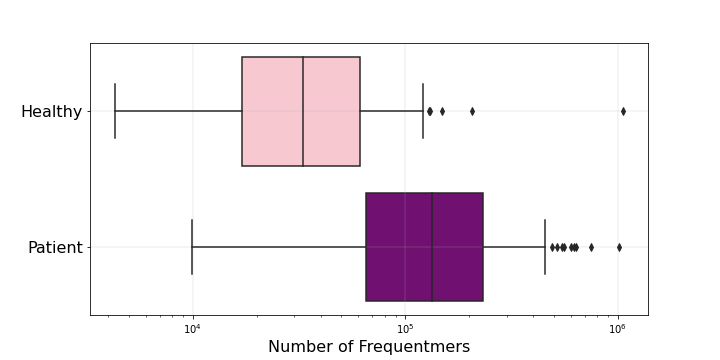

C	D

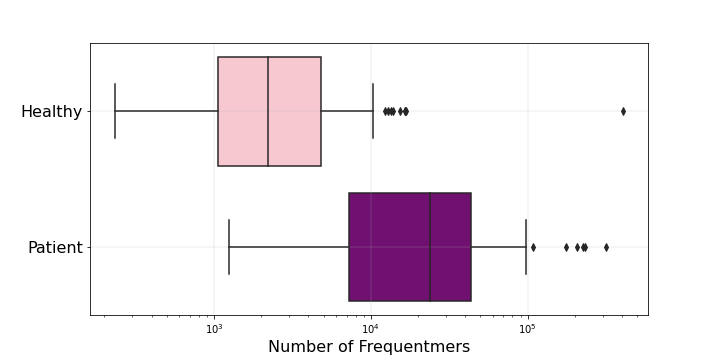

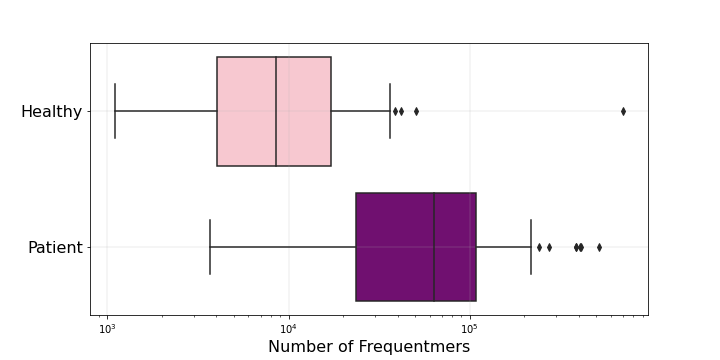

**Supplementary Figure 3: Number of frequentmers observed in the test set.** Sample recurrency of: **A.** 5, **B.** 10, **C.** 15, **D.** 20. Pink color represents healthy control frequentmers and purple represents liver cirrhosis frequentmers. All comparisons were statistically significant (Mann-Whitney U tests, p-value<0.0001).

A	B

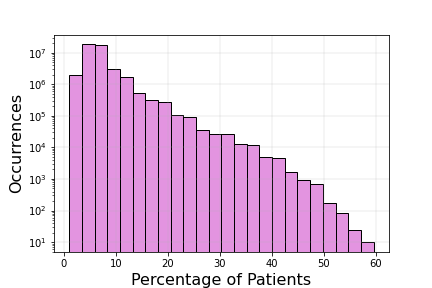

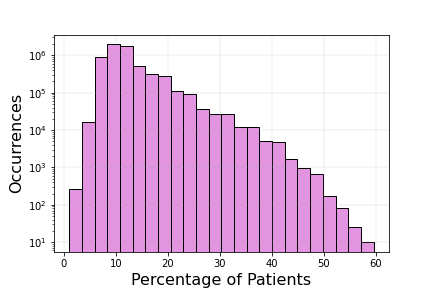

C	D

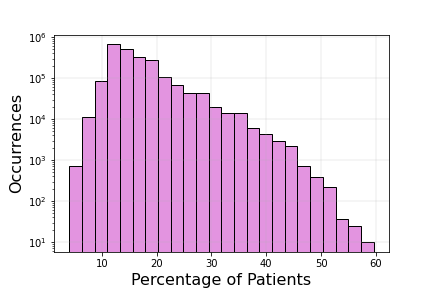

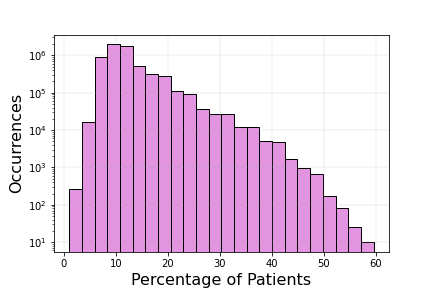

**Supplementary Figure 4: The subset of frequentmers that are only found in HBV-positive patients.** Recurrency threshold of: **A:** 5, **B.** 10, **C.** 15, **D.** 20 samples.

A	B

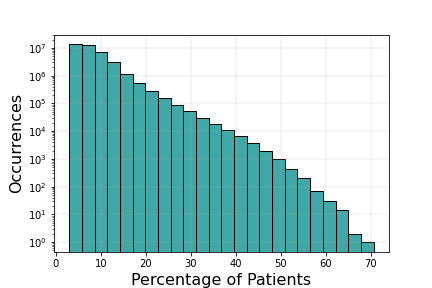

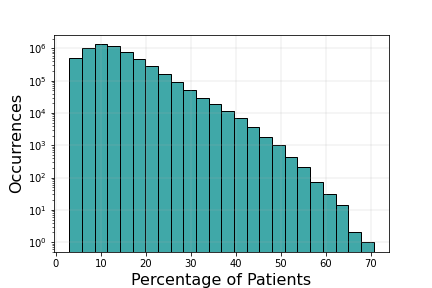

C	D

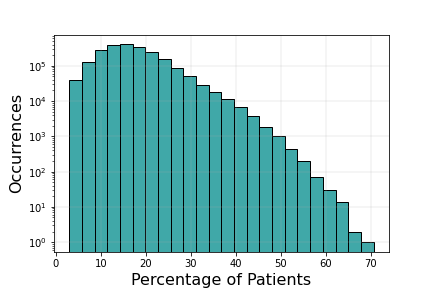

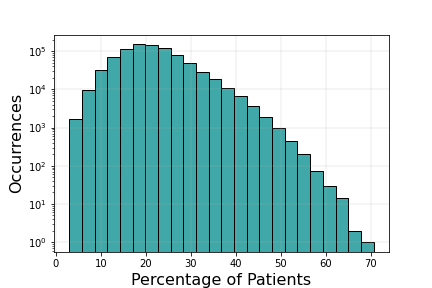

**Supplementary Figure 5: The subset of frequentmers that are only found in patients that had high alcohol intake.** Recurrency threshold of: **A:** 5, **B.** 10, **C.** 15, **D.** 20 samples.

A	B

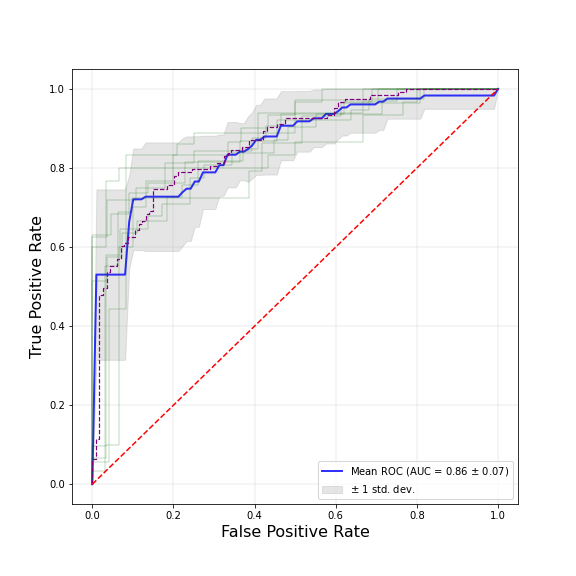

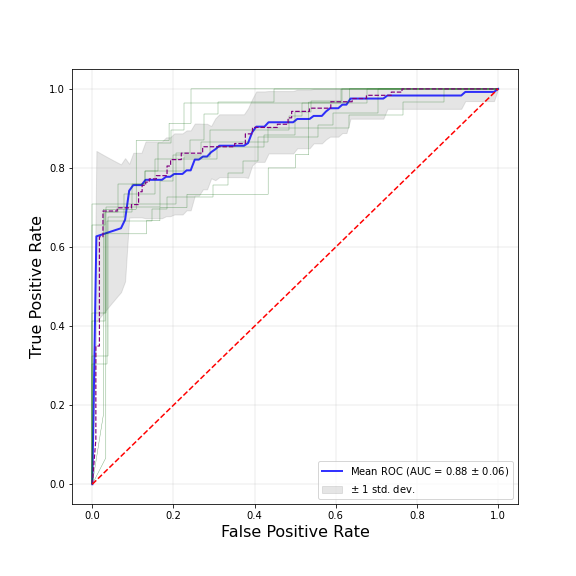

C	D

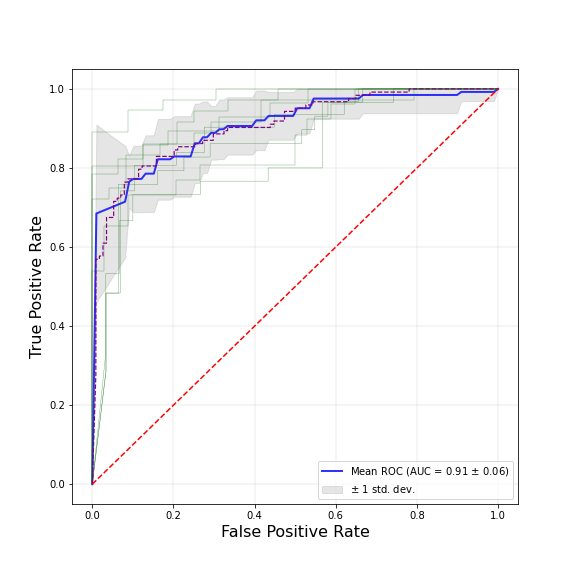

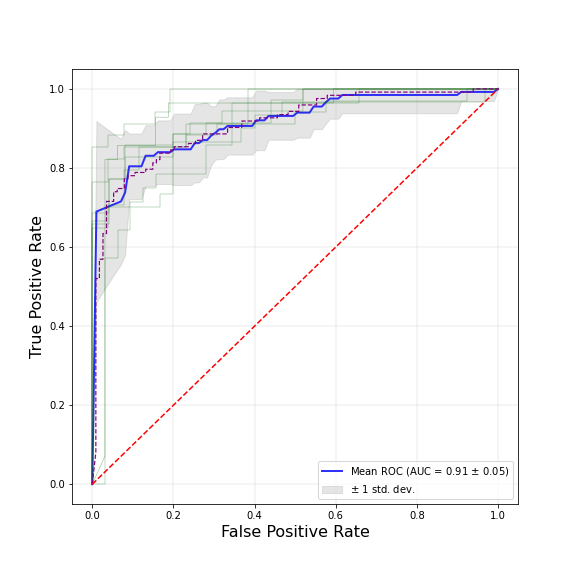

**Supplementary Figure 6: Logistic regression classification model ROC curve of liver cirrhosis and healthy control samples.** Sample recurrency of **A:** 5, **B.** 10, **C.** 15, **D.** 20. Blue line represents the mean score, green lines represent the different folds and the gray area represents confidence intervals.

A	B

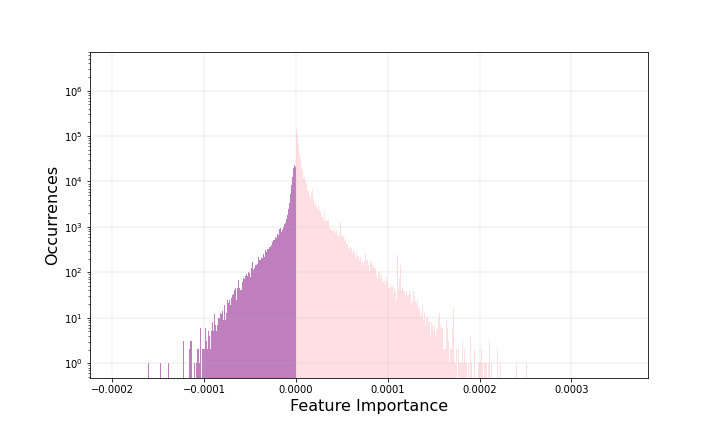

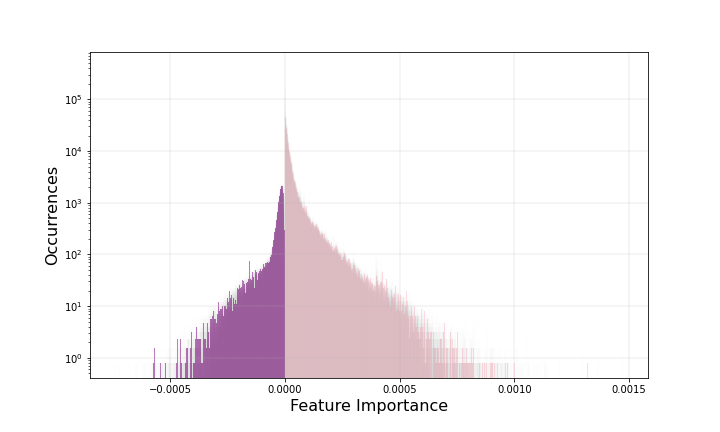

C	D

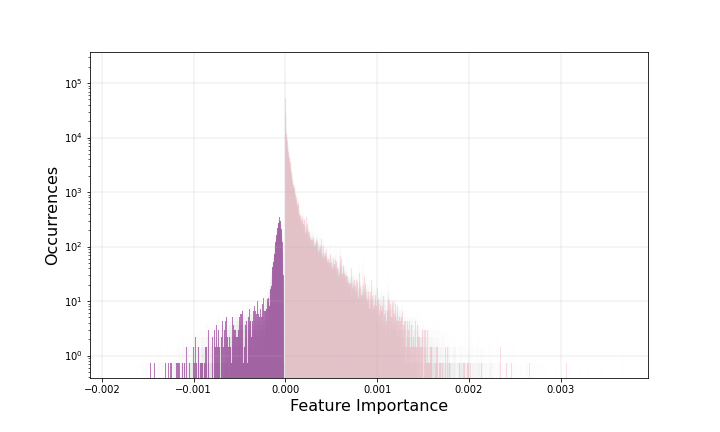

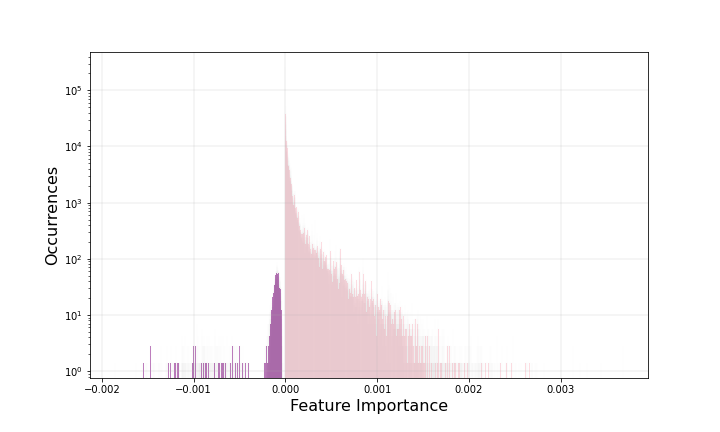

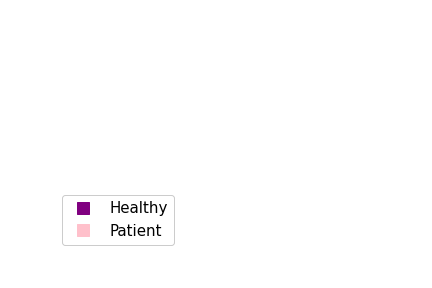

**Supplementary Figure 7: Histogram displaying the logistic regression coefficients.** Sample recurrency of: **A:** 5, **B.** 10, **C.** 15, **D.** 20.

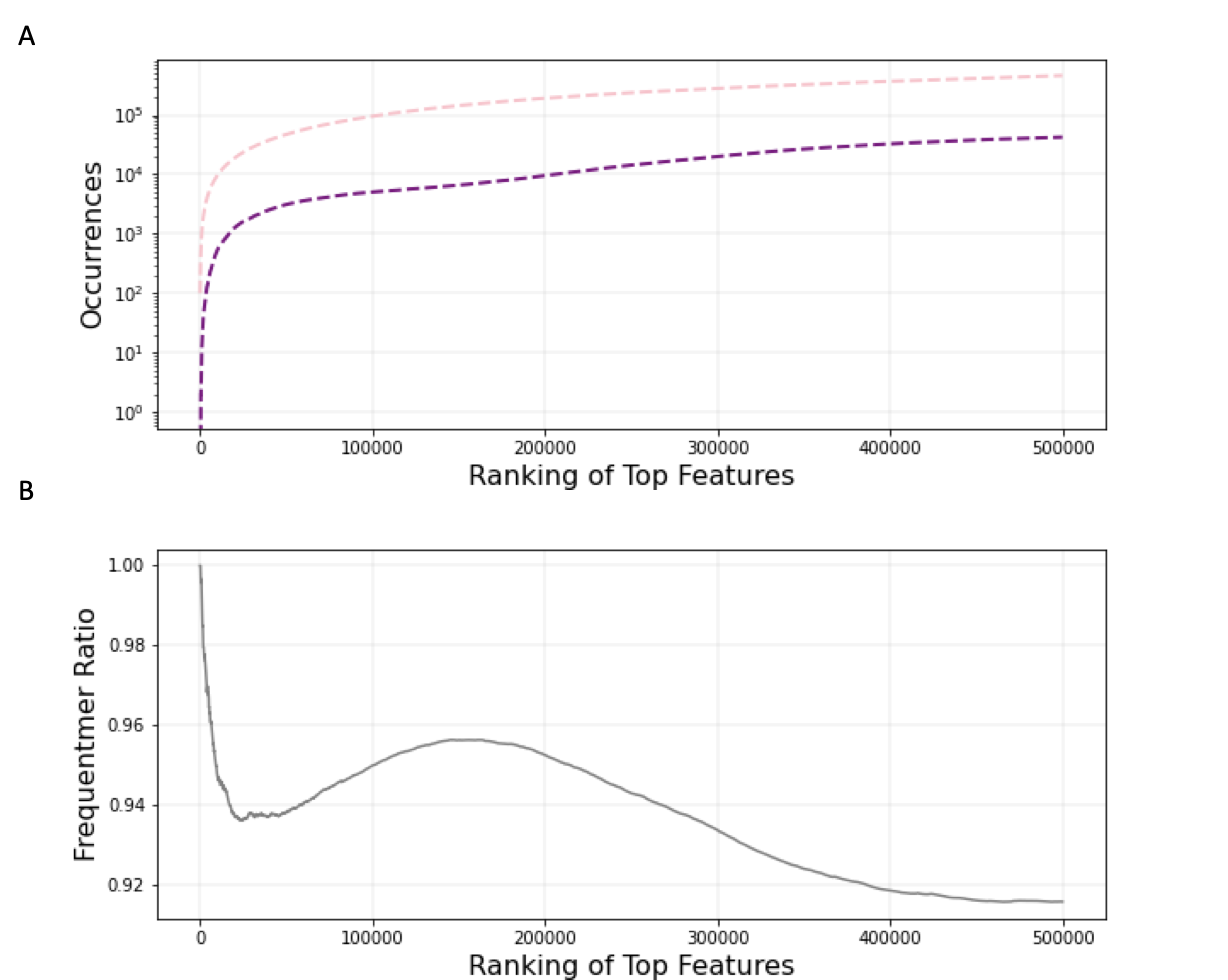

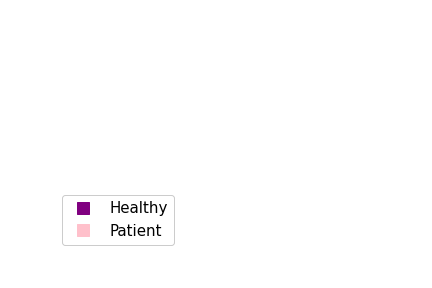

**Supplementary Figure 8: Ranked most important features by absolute coefficient score. A.** Number of most important healthy control and patient frequentmers. **B.** Frequentmer ratio for most important healthy control and patient frequentmers. Frequentmer ratio is defined as the number of patient frequentmers over total frequentmers detected. Results shown for recurrency of fifteen.

A	B

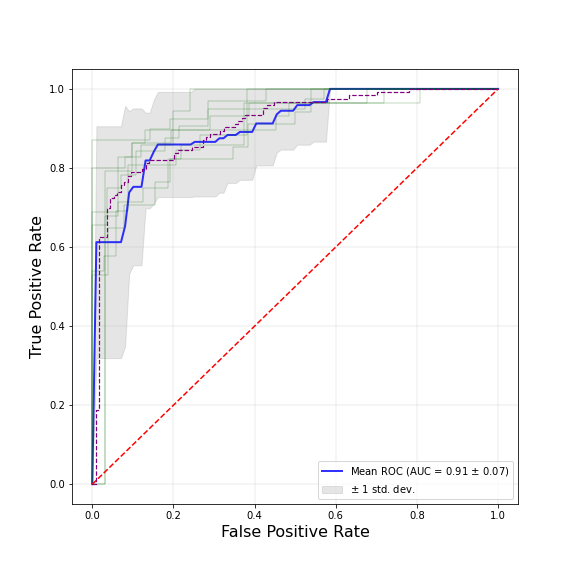

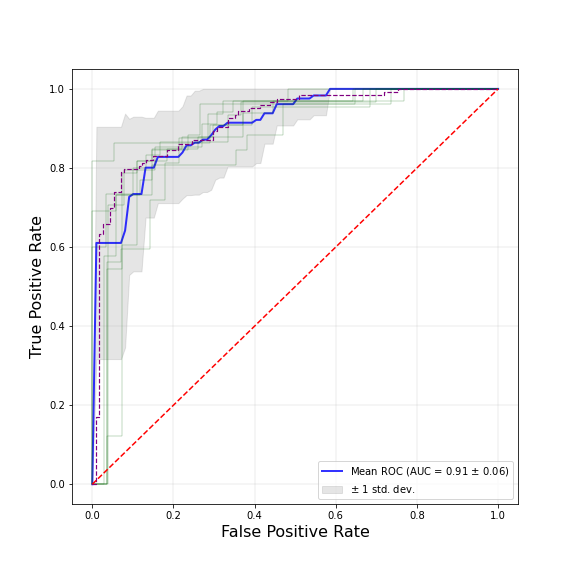

C	D

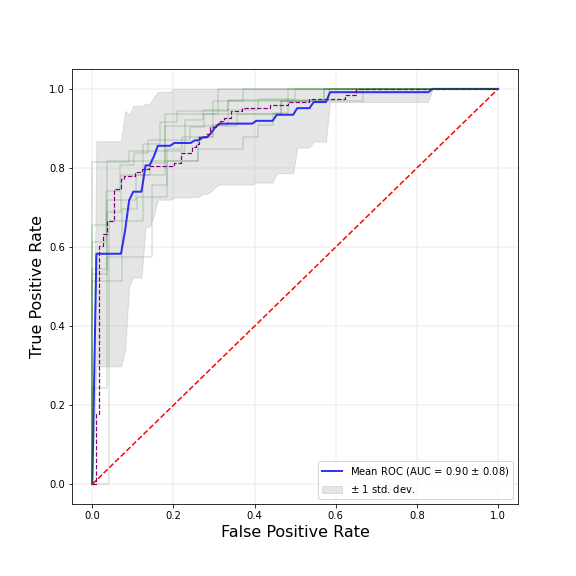

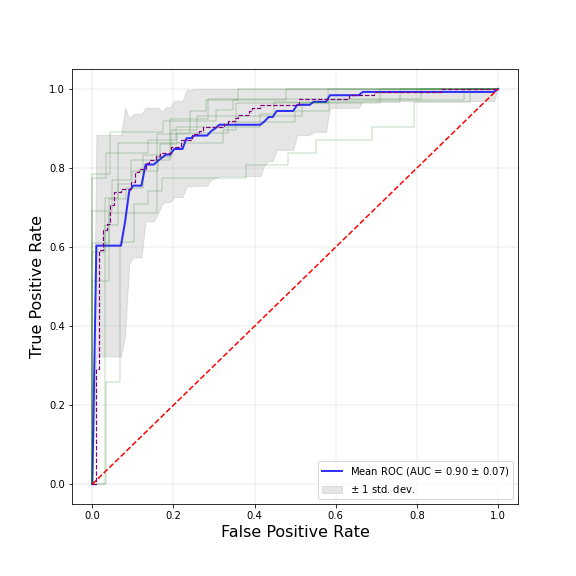

**Supplementary Figure 9: XGBoost classification model ROC curve of liver cirrhosis and healthy control samples.** Sample recurrency of **A:** 5bp, **B.** 10, **C.** 15, **D.** 20. Blue line represents the mean score, green lines represent the different folds and the gray area represents confidence intervals.
